# Influenza transmission dynamics quantified from wastewater

**DOI:** 10.1101/2023.01.23.23284894

**Authors:** Sarah Nadeau, A.J. Devaux, Claudia Bagutti, Monica Alt, Evelyn Ilg Hampe, Melanie Kraus, Eva Würfel, Katrin N. Koch, Simon Fuchs, Sarah Tschudin-Sutter, Aurélie Holschneider, Christoph Ort, Chaoran Chen, Jana S. Huisman, Timothy R. Julian, Tanja Stadler

**Affiliations:** Department of Biosystems Science and Engineering, ETH Zurich, Basel, Switzerland; Swiss Institute of Bioinformatics, Lausanne, Switzerland; Department of Environmental Microbiology, EAWAG, Dübendorf, Switzerland; State Laboratory of Basel-Stadt, Basel, Switzerland; Department of Health, Canton of Basel-Stadt, Basel, Switzerland; Cantonal Office of Public Health, Department of Economics and Health, Canton of Basel-Landschaft, Liestal, Switzerland; Division of Infectious Diseases & Hospital Epidemiology, University Hospital Basel, Switzerland / University of Basel, Switzerland

**Keywords:** Influenza, infectious disease surveillance, wastewater epidemiology

## Abstract

Influenza infections are challenging to monitor at the population level due to a high proportion of mild and asymptomatic cases and confounding of symptoms with other common circulating respiratory diseases, including COVID-19. Alternate methods capable of tracking cases outside of clinical reporting infrastructure could improve monitoring of influenza transmission dynamics. Influenza shedding into wastewater represents a promising source of information where quantification is unbiased by testing or treatment-seeking behaviors. We quantified influenza A and B virus loads from influent at Switzerland’s three largest wastewater treatment plants, serving about 12% of the Swiss population. We estimated trends in infection incidence and the effective reproductive number Re in these catchments during a 2021/22 epidemic and compared our estimates to clinical influenza surveillance data. We showed that wastewater-based incidence is better aligned with catchment-level confirmed cases than national ILI, and that only the wastewater data capture a peak in incidence in December 2021. We further estimated Re to have been below 1.05 after introduction of work from home measures in December 2021 and above 0.97 after these measures were relaxed in two out of three catchments based on wastewater data. The third catchment yielded qualitatively the same results, although with wider confidence intervals. The confirmed-case data yielded comparatively less precise estimates that include 1 before and during the period of measures. On the basis of this research we developed an online dashboard for wastewater-based influenza surveillance in Switzerland where we will continue to monitor the onset and dynamics of the 2022/23 flu season.

## Introduction

Detection and monitoring of influenza outbreaks is a crucial but challenging task. Reporting systems for influenza-like illness (ILI) and laboratory-confirmed influenza infections are used to monitor temporal trends in influenza transmission (WHO 2014), to estimate the number of symptomatic illnesses, hospitalizations, and deaths from influenza (CDC 2022b), and to help hospitals and public health officials plan vaccination campaigns and allocate treatment resources (WHO 2013). For example, doctors may prescribe influenza-specific treatment based on knowledge of an ongoing influenza outbreak in the region and a symptomatic diagnosis, before waiting for laboratory confirmation (WHO 2022a). This underscores the public health relevance of accurate detection and monitoring of influenza outbreaks.

Existing influenza surveillance systems based on clinical data have been severely impacted by the COVID-19 pandemic. First, many symptoms of COVID-19 and influenza overlap, which complicates symptom-based influenza diagnosis (CDC 2022a). Test-seeking behavior also changed during the pandemic, reportedly increasing compared to pre-pandemic in the U.S. (CDC 2022b). Finally, pandemic-related public health measures and associated behavioral changes have disrupted typical seasonal influenza transmission dynamics (Dhanasekaran et al. 2022; Boehm et al. 2022). Thus, pandemic-related changes have simultaneously changed influenza transmission dynamics and decreased the reliability of ongoing influenza surveillance efforts. Many influenza surveillance reports accordingly include COVID-related disclaimers about the representativeness and interpretability of the data (CDC 2022b; FOPH 2022b; WHO 2022b).

Wastewater surveillance represents a promising alternative method for pathogen surveillance that is independent of testing behavior, capable of discriminating between diseases with overlapping symptoms, and can capture unreported cases (Fernandez-Cassi et al. 2021). Many pathogens are shed by infected individuals via stool, sputum, and/or from the skin, depending on the pathogen.This means pathogen particles can make it into wastewater when infected individuals use the toilet, brush their teeth, or shower. Previous studies have shown that a range of pathogens are detectable in wastewater samples (Xagoraraki and O’Brien 2020; Kilaru et al. 2022). In regions where household wastewater is centrally collected, wastewater represents a pooled community sample and wastewater pathogen loads indicate community disease burden. The idea of wastewater-based epidemiology is not new, but has recently experienced a surge in popularity, with many communities developing detection and monitoring strategies for SARS-CoV-2 in wastewater (Medema et al. 2020).

Wastewater-based surveillance offers the opportunity to better understand influenza dynamics, similar to its role in understanding COVID-19 dynamics. Wolfe et al. (2022) established that influenza A virus (IAV) in wastewater correlated well with incidence rates from two well-characterized outbreaks on U.S. university campuses. Mercier et al. (2022) similarly quantified IAV in wastewater at the municipality scale in Ottawa, Canada. They established that IAV concentrations in wastewater correlated well with municipal surveillance data, with the wastewater data evidently leading clinical surveillance data by 17 days. They also extended wastewater surveillance by sub-typing the detected IAV. Most recently, Boehm et al. (2022) developed a multiplexed method to quantify influenza A and B alongside several other respiratory viruses in wastewater. They confirmed that IAV concentrations in wastewater at the municipal level were associated with laboratory-confirmed cases at the state level. Influenza B virus (IBV) concentrations were low and frequently non-detectable. These previous studies indicate IAV is detectable in wastewater and can be used to study transmission dynamics in the community.

In this study, we aimed to implement wastewater surveillance for influenza at the national scale in Switzerland and to estimate IAV and IBV transmission dynamics from wastewater. To do this, we measured the concentration of seasonal influenza types A and B at the three largest wastewater treatment facilities in Switzerland from December 2021 to April 2022. Combined, these facilities serve approximately 12% of the Swiss population. Following a previously established method, we deconvoluted wastewater influenza loads to estimate trends in infection incidence and the effective reproductive number (Re). We compared the wastewater-based results with laboratory-confirmed infection incidence in each catchment area. We continue to measure IAV and IBV concentrations in these catchments since October 2022. The results presented here, as well as the results of our ongoing monitoring efforts, are available on an online dashboard at https://influenza.wastewater.science/. All the analysis and dashboard code is open-source. We anticipate these results and resources will help inform public health officials and hospitals about the onset and intensity of future influenza seasons.

## Methods

### Sampling RNA in wastewater

RNA was extracted for IAV and IBV quantification from wastewater samples collected at Swiss wastewater treatment facilities in Zurich, Geneva, and Basel. The respective catchment areas served by these facilities are shown in Figure 1.

**Figure 1.**
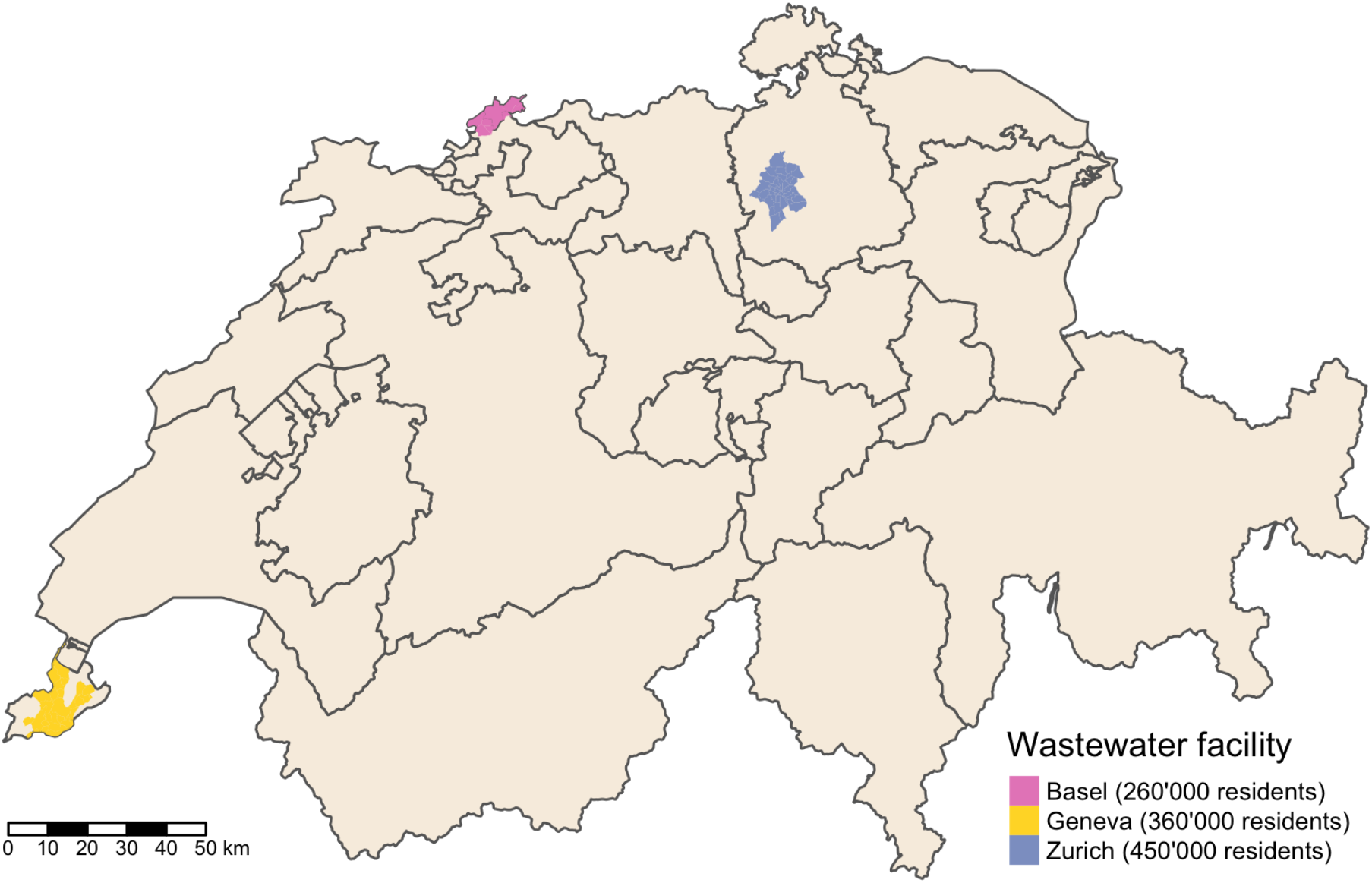
Catchment area map. Colored areas show the catchment areas for the three wastewater treatment facilities where influenza A and B virus loads were measured. The Basel facility serves several communities in France (Neuwiller) and Germany (Weil-Otterbach and Inzlingen) that are not shown. Also not shown is the community of Brüglingen in Münchenstein, which is served by the Basel facility.

For the Zurich and Geneva facilities, 24-hour flow-composite raw influent wastewater samples were collected within the scope of ongoing wastewater-based SARS-CoV-2 surveillance. Total nucleic acid was extracted from 40 ml of raw influent using a modified version of the Wizard(R) Enviro Total Nucleic Acid Kit (Promega Corporation, Madison, USA). Nucleic acids were eluted in 80 μl of RNAse/DNAse-free water, and further purified using a OneStep PCR Inhibitor Removal Kit (Zymo Research, Irvine, CA, USA). Nucleic acid extracts were stored at -80C for up to a year prior to analysis.

For the Basel facility, 24 hours composite samples were collected daily (except for two days, generally on weekends, when 48 hours composite samples were taken) from the inflow of ProRheno AG, Basel, the local wastewater treatment plant (Bagutti et al. 2022). Until further processing, samples were stored at 4° C for a maximum of 72 hours. Total nucleic acid was concentrated and extracted from 40 ml of wastewater using the Maxwell^®^ RSC Environ Wastewater TNA Kit (Promega, Madison, USA). Nucleic acid extracts were stored at -20° C for up to two weeks and at -80° C for up to a year prior to analysis.

### Quantifying influenza RNA

For the Zurich and Geneva facilities, we quantified IAV and IBV from at least two replicate aliquots from on average two samples per week from 1. December 2021 to 30. April 2022. Quantification was done using digital reverse transcription PCR (RT-dPCR) on the Crystal Digital PCR Naica System (Stilla Technologies, France) with qScript XLT One-Step RT-qPCR Kit (CN: 95132-02K, Quantabio, MA, USA). Two assays were used, IABV and RESPV4. IABV is a duplex assay with IAV and IBV, targeting the matrix proteins of each respective virus. (Table S1). RESPV4 is an updated tetraplex assay that incorporates SARS-CoV-2 Nucleoprotein locus 2 (N2) and Respiratory Syncytial Virus Matrix protein (RSV), while including the aforementioned IAV and IBV targets. (Table S1). The mastermix final volume was 27 μL, with 5.4 μl of template and 21.6 μL of pre-mix. The premix consists of the US CDC SARS-CoV-2 RUO Kit N2, with a primer and probe final concentration of 500 nM and 125 nM, respectively (Integrated DNA Technologies, n.d.) (Integrated DNA Technologies, USA). IAV, IBV and RSV all had primers at a final concentration of 500 nM The RSV probe had a final concentration of 400 nM, and the IAV and IBV probes were at a final concentration of 200 nM. Fluorescein was added at a final concentration of 125 nM as a background dye for droplet detection. Finally, RNAse/DNAse-free water was added (variable, depending on stock concentrations) to bring the pre-mix to the correct final volume, of which 25 μL was pipetted into Sapphire Chips (Stilla Technologies). Thermocycling conditions included droplet partitioning at 40°C for 12 minutes, followed by reverse transcription at 50°C for 1 hour, and then followed by 40 cycles of 95°C for 30s and 57.5°C for 60s. Chips were subsequently read on the Stilla Prism3 chip reader for the IABV assay or the Stilla Prism6 chip reader for the RESPV4 assay.

Analysis of IAV included results from both IABV and RESPV4, whereas with IBV only the measurements for RESPV4 were used due to insufficient separation of positive droplets (Figure S1, S2). Samples were run in technical duplicate, and each measurement consisted of 23’648 droplets on average, with a standard deviation of 6628 droplets. Each droplet is assumed to have a volume of 519 nL. Measurements were only included if they had at least 15’000 droplets. Droplets of incorrect volume (polydispersity) if present were manually excluded from the analysis. Quality control included one no-template control and one positive control with every five samples, and inhibition testing for the SARS-CoV-2 N1 locus for every sample as previously described.

For the Basel facility, we quantified IAV and IBV in single aliquots from on average two samples per week. The number of IAV and IBV was determined in a triplex one-step RT-qPCR reaction using 5 μl wastewater extract with the GoTaq^®^ Enviro FluA/FluB /SARS-CoV-2 System (Promega) and the protocol provided in the manual.

We converted the viral concentration measurements (in genome copies per mL) to daily pathogen loads (genome copies per day) by multiplying the concentration measurements by total inflow to the respective wastewater treatment facility on the sample date.

### Estimating trends in influenza incidence from wastewater

We used our previously developed approach to estimate trends in infection incidence and the effective reproductive number from pathogen data in wastewater (Huisman et al. 2022). The actual estimation was done using the R package estimateR (Scire et al. 2022).

First, we scaled the wastewater load data by the treatment plant-specific minimum detected load (1.6×10^11^ for Zurich, 1.1×10^11^ for Geneva, and 3×10^9^ for Basel). This corresponds to the assumption that the minimum detected load represents one infected person. Such an assumption is necessary to make the range of wastewater measurements comparable to incidence in an outbreak, since the deconvolution method is optimized for case data and does not perform well when inputs are orders of magnitude higher (as with wastewater loads) (Huisman et al. 2022). As a sensitivity analysis, we also tried using 10x,100x, and 1000x more scaling, i.e. the minimum detected viral load corresponds to 10, 100, or 1000 infected individuals in the catchment rather than 1. Next, we performed linear interpolation to generate a regular daily time series of measurements. We chose linear rather than cubic spline interpolation because although the latter generated more realistic smooth changes during most periods, it produced spurious lows where wastewater measurements oscillated dramatically during a few short time periods. Finally, we used LOESS smoothing to smooth the time series. In both the interpolation and smoothing steps, we make the implicit assumption that the true influenza incidence in a catchment does not change drastically day-to-day and that instead, temporal noise in wastewater measurements comes from other factors like variation in laboratory processing and detection methods, differing residence time in the sewers depending on the source, biofilm sloughing, or stochastic noise associated with temporally varying inputs (Wade et al. 2022; Zahedi et al. 2021).

As in Huisman et al. (2022), we used a pathogen shedding load distribution to deconvolve the wastewater load data to estimate trends in infection incidence. This distribution describes the relative amount of virus an infectious individual sheds into the wastewater every day after infection. We used two different shedding load distributions, one based on influenza virus load measured in respiratory samples (Carrat et al. 2008) and one based on influenza virus load measured in fecal samples (Hirose et al. 2016; Chan et al. 2011). We fit a gamma distribution to the measurements for each sample type given in the respective publications (see Supplemental methods). The resulting respiratory shedding load distribution has a mean of 2.5 days from infection and a standard deviation of 0.9 days and the fecal shedding load distribution has a mean of 12.2 days from infection and a standard deviation of 7.6 days (Figure S3). The fecal shedding load distribution includes a fixed two-day shift to account for the fact that measurements are referenced from the time of symptom onset and thus neglects shedding prior to symptom onset (Supplemental methods). We used this distribution as a sensitivity analysis and present results in the main text using the respiratory shedding load distribution. We assumed delays inside the sewer system to reach the wastewater sample collection point are negligible. Finally, we quantified uncertainty in the infection incidence deconvolution by performing 500 block-bootstrap replicates of the measurement error, as described in Huisman et al. (2022).

### Estimating the effective reproductive number

We also quantified the effective reproductive number (Re) through time using estimateR. Re is the expected number of secondary infections generated by a single infectious individual at a certain time point. This metric is commonly used for monitoring disease transmission dynamics, in particular because there is an easily interpretable threshold at Re = 1 that indicates whether an outbreak is expected to grow (Re > 1) or decline (Re < 1). Re estimates also put wastewater- and case-based metrics on a common scale. Re can vary through time according to population immunity, infection control measures, and behavioral changes.

We used the method of Cori et al. (Cori et al. 2013) implemented in estimateR to generate an Re trajectory estimate from each bootstrapped replicate for infection incidence. For this, we used an influenza-specific serial interval with a mean of 2.6 days and a standard deviation of 1.5 days (Ferguson et al. 2005) and a sliding window of 3 days (the software default). We combined the uncertainty in the Re estimation, which is reported as the 95% credible interval for Re, with the uncertainty across the bootstrap replicates for infection incidence, which is reported as the 95% bootstrap confidence interval (estimateR option “combine_bootstrap_and_estimation_uncertainties”). By using the union of the highest of each type of uncertainty, we account for both estimate uncertainty and measurement uncertainty.

### Comparison to clinical influenza data

Two types of clinical data on influenza are collected by the Federal Office of Public Health (FOPH) in Switzerland. These are consultations for ILI in the sentinel system “Sentinella” reported voluntarily by participating healthcare providers and laboratory-confirmed cases that are mandatorily reported by diagnostic laboratories (FOPH 2022b). We downloaded national ILI incidence from the FOPH influenza report (FOPH 2022b). We received the number of laboratory-confirmed cases from the FOPH stratified by week, influenza type, and postal code from 24. August 2021 onwards for the catchment areas of several Swiss wastewater treatment plants. To generate an estimate for the number of cases specific to each sampled catchment area, we multiplied the number of cases in each postal code by the percentage of residents contributing to the relevant catchment, based on a delineation of the catchment boundary, and then summed across all contributing postal codes. We note that confirmed cases were also reported outside of our study period, in this text we show only data from 1. December 2021 to 30. April 2022.

We generated Re estimates from the catchment-level confirmed case data using the same procedure as for the wastewater data, except with cubic spline (rather than linear) interpolation to estimate daily cases from the weekly reported data. We also used a delay distribution based on a timeline of symptom severity for the case data rather than viral shedding. This is based on the assumption that the probability a person seeks a test is linked to their symptom severity, with most individuals getting tested on the day of their peak symptoms. We generated this distribution by fitting a gamma distribution to data from Carrat et al. (2008), resulting in a distribution with a mean of 4.5 days from infection and a standard deviation of 0.8 days (Supplemental methods, Figure S3).

## Results

During the period from 1. December 2021 to 30. April 2022 we were able to detect IAV in wastewater on more than 90% of sampled days (IAV detected on 37/38 days from Zurich, 39/42 days from Geneva, and 45/50 days from Basel). IBV was less often detectable in wastewater (IBV detected on 7/35 days from Zurich, 9/33 days from Geneva, and 1/50 days from Basel). Both the wastewater load data and weekly confirmed case data suggest one or more IAV outbreak peaks each in each catchment area (Figure 2A). However, only an IAV peak occurring roughly from March - May 2022 is robustly supported across all three catchments by both the wastewater and confirmed case data. Finally, we observe that the catchment-level confirmed case data corresponds better with wastewater measurements than national ILI, which reached a peak in late January 2022 that was similar in size to a later peak in mid-March 2022 (Figure 2).

**Figure 2.**
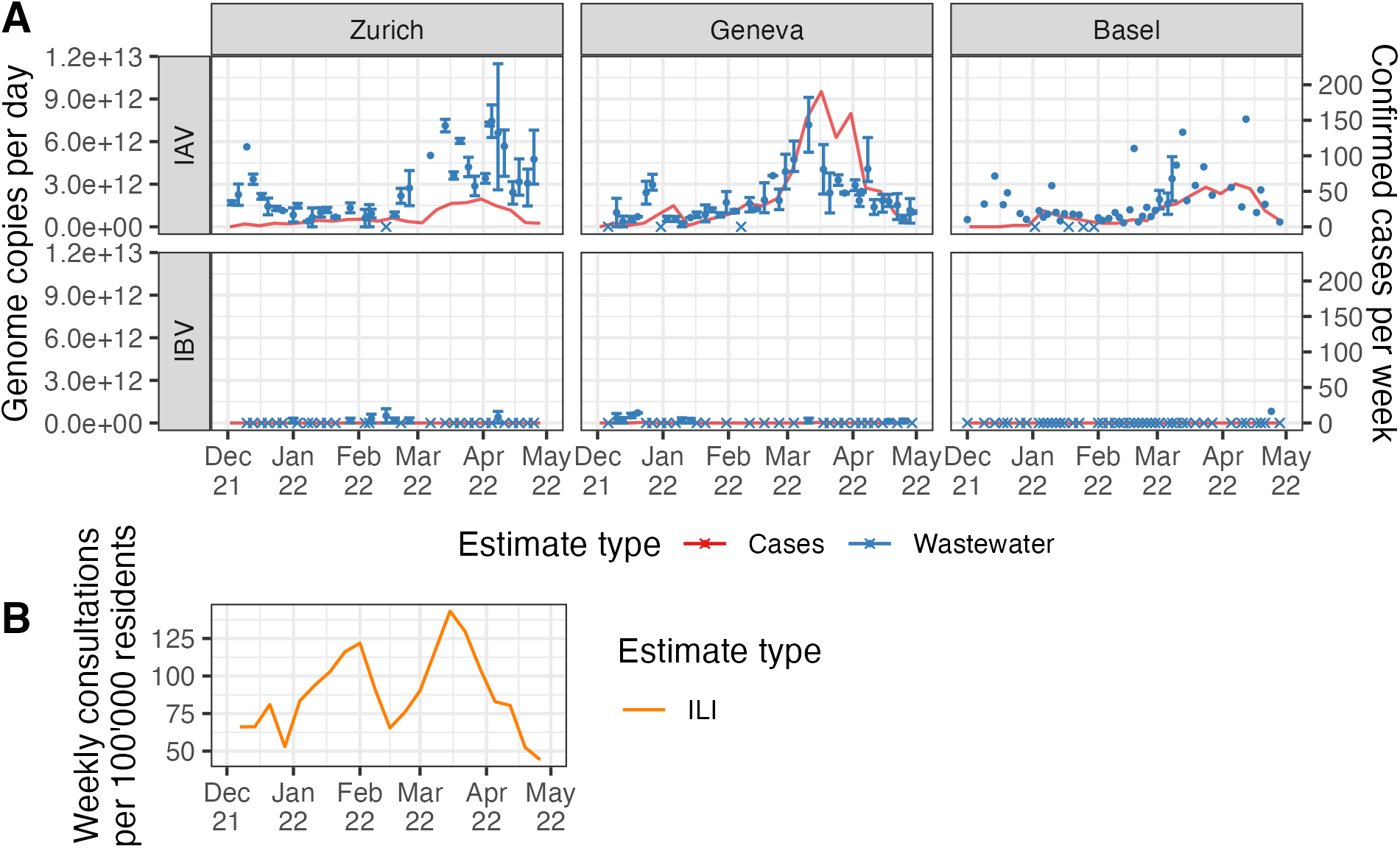
Influenza measurements from wastewater versus clinical data. **(A)** shows the two data sources used to estimate Re. Wastewater measurements (in blue) are shown as the mean (points) and range (error bars) of measurements across aliquots from each sampled day. Days where no aliquots had detectable virus are shown with crossed rather than round points. Note that wastewater quantification methods differed between the Zurich and Geneva catchments and the Basel catchment. Confirmed cases (in red) are shown as a line connecting weekly reported case numbers within the catchment. **(B)** shows influenza-like illness data (in orange) reported nationally during the same time period for comparison.

We can compare catchment-specific influenza outbreak dynamics based on wastewater data versus laboratory-confirmed case data by deconvolving both the wastewater loads and case data to estimate trends in infection incidence. By applying the appropriate delay distribution to each type of data, we can line up incidence estimates on a comparable time scale (Figure 3). There are two points worth highlighting in this analysis. First, the magnitude of wastewater-based infection incidence estimates are sensitive to the scaling of wastewater loads. Our scaling based on the assumption that the minimum detected load corresponds to a single infected individual should be conservative, meaning infection incidence is probably higher than reported here. This is because we assume the minimum correspondence between wastewater loads and infections in the catchment (the minimum detected wastewater load corresponds to a single infected individual). Second, since there were very few laboratory-confirmed cases of IBV during the study period, we only show case-based results for IAV. There were 6 IBV confirmed cases in the Geneva catchment and none in the Zurich or Basel catchments. In comparison, there were 965 IAV confirmed cases in the Geneva catchment, 359 in the Basel catchment, and 255 in the Zurich catchment (Figure S4). Similarly, we do not report results for IBV in Basel wastewater, since it was only detected on one day.

**Figure 3.**
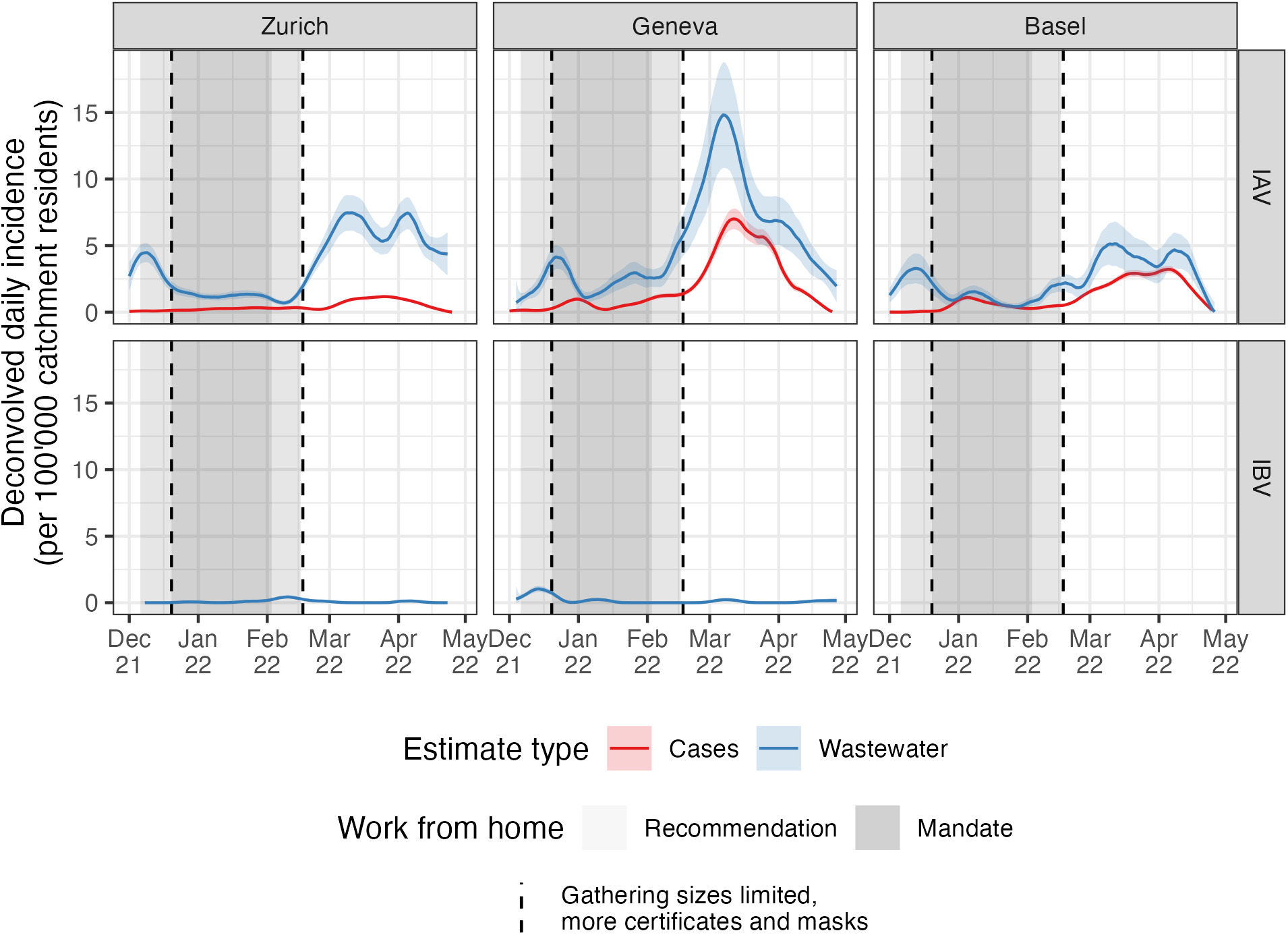
Trends in infection incidence. Infection incidence estimates are based on a deconvolution from wastewater influenza measurements (blue) or from laboratory-confirmed influenza A cases in each catchment (red). Influenza B case numbers were too low to run the estimateR pipeline on. Shaded areas show when work from home measures were in place and dashed lines show the start and end dates of stronger measures to limit gathering sizes and require COVID certificates and masks in more situations to combat the Omicron variant of SARS-CoV-2 (see Table 1).

The estimated trends in infection incidence deduced from the wastewater data show strong evidence of an IAV outbreak in Zurich, Geneva, and Basel in December 2021; this outbreak is not observed in the confirmed case data (Figure 3). The same trend is evident in the raw wastewater load data as well (Figure 2). This outbreak observed in wastewater appears to have peaked around the same time measures were introduced to reduce population mobility and contacts in Switzerland in order to combat the spread of the Omicron variant of the SARS-CoV-2 virus (Table 1). Around the time measures were lifted (first half of February), IAV incidence again increased in all three catchments according to wastewater-based estimates. Confirmed IAV infections increased during this period as well.

**Table 1.** Selected measures to combat the Omicron variant of SARS-CoV-2 in Switzerland. The table only shows the major measures for reducing population mobility and contacts, a complete list of measures is available at (FOPH 2022a).

| Start date | End date | Measure | Source |
| --- | --- | --- | --- |
| 6. Dec 2021 | 17. Feb 2022 | Work from home recommendation | FOPH 2022a; SWI 2022 |
| 20. Dec 2021 | 3. Feb 2022 | Work from home mandate | FOPH 2022a |
| 20. Dec 2021 | 17. Feb 2022 | Gathering sizes limited;<br>COVID certificates and masks required in more situations | SWI 2022 |
| 17. Feb 2022 | 30. Mar 2022 | None except isolation of people with a positive test and masks in public transit and health care settings | Federal Council 2022 |

IBV incidence was estimated to be much lower than IAV incidence in the Zurich and Geneva catchments, matching the observation of many fewer confirmed IBV cases than IAV cases. There is very weak support for a small peak in IBV infections in Geneva around the time mobility restrictions were introduced and in Zurich around the time they were lifted based on the wastewater data. However, these results are based on detection of IBV at low concentration in only a few samples on a handful of days (Figure 2, S1).

Generally, we note that depending on the shedding load distribution assumed, results on infection incidence for both IAV and IBV shift in time. Given a shedding load distribution based on potentially longer fecal shedding rather than respiratory shedding as shown here, all estimates shift 1-2 weeks further to the past (Figure S5). However, a qualitative correspondence between the wastewater-based estimates and mobility restriction measures remains.

Next, we used the effective reproductive number (Re) to get further insight into the epidemic dynamics. Re helps us identify when influenza outbreaks were growing or declining with certainty (when confidence bounds on Re exclude the epidemic threshold of 1). We note that Re estimates are comparably robust to different scalings of the wastewater load data, at least up to several order of magnitude differences, since these estimates are only based on relative changes in incidence over time (Figure S6). As for the incidence estimates, the timing of the Re estimates depends on the chosen shedding load distribution (Figure S6).

Figure 4 shows that for IAV in Zurich and Geneva, the wastewater-based Re dropped below 1.05 upon introduction of mobility restriction measures to combat the Omicron variant. The same decreasing trend is observed in the wastewater-based Re from Basel, although confidence intervals are wider around the epidemic threshold. Later Re was above 0.97 in the Zurich and Geneva catchments after relaxation of the measures. We note that assuming less conservative scalings for wastewater loads would only increase the certainty of these Re estimates, pushing the lowest values significantly below 1 and the highest values significantly above 1 (Figure S6). Confirmed cases of IAV in each catchment were also very low for most of the sample period. This is responsible for the delayed start of Re estimates and large confidence bounds for case-based estimates in Figure 4. We also have confirmed case data for postal codes corresponding to several other catchment areas where wastewater surveillance was not yet done. Combining all confirmed cases from all postal codes for which we have data yields an Re estimate indicating Re was above 1 in Switzerland in early March 2022 (Figure S7). However, even combining all available case data, case-based Re estimates are still too uncertain in December 2021 to draw any conclusions. Thus, the wastewater-based Re estimates are more precise than confirmed-case based estimates, at least at the catchment scale.

**Figure 4.**
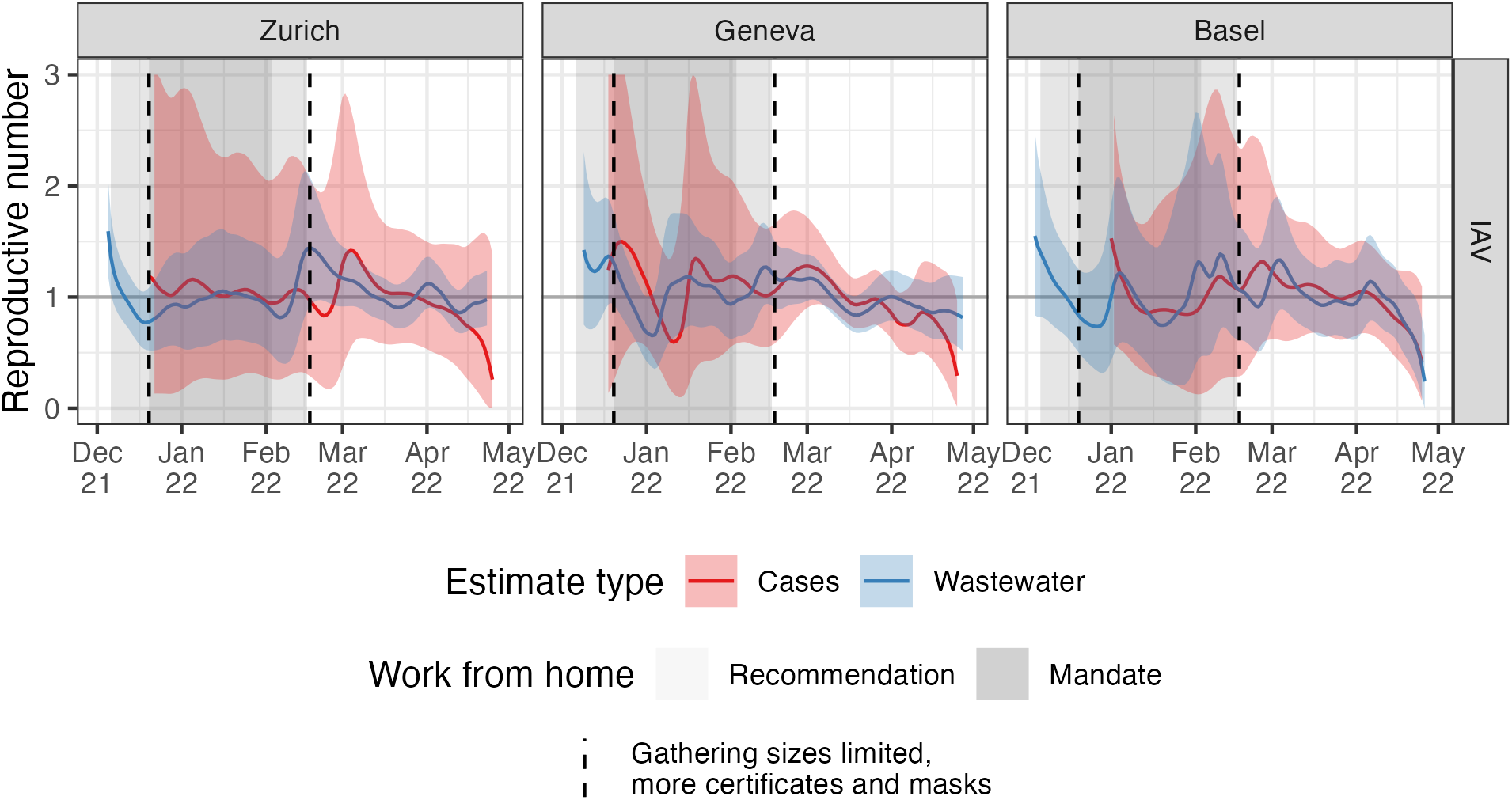
Reproductive number estimates. The different colors show estimates based on wastewater influenza measurements (blue) and laboratory-confirmed influenza cases (red). Shaded areas show when work from home measures were in place (see Table 1).

Incidence of IBV based on wastewater and confirmed cases was too low to generate reliable Re estimates (Figure S7).

All the results shown here are also made available on an online dashboard at https://influenza.wastewater.science/. We will continue to update this dashboard with ongoing surveillance results.

## Discussion

In this work, we presented proof-of-concept results for quantification of influenza transmission dynamics based on wastewater viral loads. We were able to estimate trends in infection incidence and quantify the effective reproductive number for IAV in the three largest wastewater catchment areas in Switzerland. IBV was occasionally detectable in these catchment areas but at low concentration. Combined, these data paint a different picture of influenza outbreak dynamics compared to confirmed case data, with the wastewater-based dynamics qualitatively aligning better to population mobility restrictions in winter 2021/22 due to the Omicron variant of SARS-CoV-2. Moving forward, we plan to expand our wastewater-based surveillance project to include additional catchment areas for which we have laboratory-confirmed case data (Figure S4). This will enable further validation of the trends observed in this proof-of-concept project.

A primary limitation of this study, and indeed wastewater-based pathogen surveillance in general, is that pathogen shedding, transport through the sewershed, and decay dynamics in wastewater are poorly understood. Depending on the specific pathogen and sewershed characteristics, these dynamics likely vary between pathogen types and wastewater catchments. For our specific Re estimation method, we rescaled wastewater loads to incidence values using the minimum detected wastewater viral load in each catchment. This scaling depends both on the detection limit of our quantification method and the specific sewershed. Thus, we cannot compare the absolute magnitudes between the wastewater- and confirmed case-based incidence estimates shown in Figure 3, nor can we compare these magnitudes for wastewater-based incidence across catchments. However, the relative values should be comparable across influenza types (IAV and IBV) within the same catchment. Re measurements, on the other hand, are comparable across catchments and pathogen types since estimates are based on relative changes through time.

We performed several sensitivity analyses to test the robustness of our results to unknown influenza dynamics in wastewater. First, we showed that Re measurements are robust to a several orders of magnitude difference in scaling of wastewater loads, except that confidence bounds become arbitrarily small when the scaling results in higher incidence estimates (Figure S6). The scaling we used gives comparably conservative (wide) confidence bounds. Then, we also performed a sensitivity analysis for the shedding load distribution, taking into account fecal versus respiratory influenza shedding. This showed that the magnitude of our incidence and Re estimates are generally robust to unknown shedding dynamics but that estimates may be shifted too far towards the present, depending on whether potentially longer fecal shedding is really the major driver of wastewater influenza loads (Figure S5). We justify the use of a respiratory shedding load distribution here by noting that respiratory shedding appears to be orders of magnitude greater than fecal shedding (Figure S8) (Chan et al. 2011; Hirose et al. 2016; Lau et al. 2010; To et al. 2010). We also note that trends in wastewater-based infection incidence align better with confirmed-case based estimates when assuming a respiratory shedding load distribution (Figure 3, S5). However, the timeline of these case-based estimates are also subject to our assumption of a delay distribution based on symptom severity over time. As a result, we cannot make any concrete statements on the lead or lag times between wastewater and case-based indicators of influenza incidence. We plan to update the shedding load and time-to-case-confirmation distributions used in our accompanying dashboard should more data become available.

This project highlights the potential of wastewater-based surveillance for generating public health-relevant insights beyond SARS-CoV-2. First, we showed that confirmed case data likely better represented influenza incidence during the 2021/22 season than ILI data based on a correspondence with our wastewater measurements. Then, we showed that wastewater measurements were more sensitive to a peak in IAV incidence in Switzerland in December 2021 and yielded more precise Re estimates compared to confirmed case data on the same geographic scale.

To our knowledge, this is the first time a mechanistic model has been applied to quantify influenza transmission dynamics from wastewater data. We emphasize that the development of mechanistic models for wastewater-based epidemiology is still in its infancy. Such models could in principle incorporate both clinical and wastewater data at the same time, and more detailed assumptions on noise generating processes. Methodological developments along these lines should improve the robustness and precision of wastewater-based estimates for pathogen transmission dynamics.

However, a prerequisite for application of complicated mechanistic models is a robust pathogen quantification method for wastewater. Our results demonstrate that both RT-qPCR and dRT-PCR based quantification methods are capable of sufficiently sensitive detection to reconstruct estimated infection incidence and estimate Re. In particular we plan to extend the dPCR assay to detect more pathogens as Boehm et al. did in the U.S. (Boehm et al. 2022).

We are continuing to measure IAV and IBV loads in Swiss wastewater since October 2022 and generate corresponding estimates for infection incidence and the effective reproductive number by catchment area. These results are available on an online dashboard at https://influenza.wastewater.science/. We make all the code for the analysis and this dashboard available on our project repository at https://github.com/wise-ch/wastewater-influenza-dashboard. We envision these results will help improve influenza surveillance in Switzerland by serving as an alternate source of information that is less susceptible to some case-based surveillance challenges amplified by the COVID-19 pandemic. More generally, we envision that our surveillance dashboard and the open-source code supporting it can serve as a blueprint for international surveillance efforts. Finally, as we generate more seasons of high-quality wastewater data and develop detailed mechanistic models, we expect we can move beyond surveillance to generate new insights on the drivers of influenza transmission based on wastewater data.

## Data Availability

The catchment-level wastewater load and confirmed case data used in this manuscript is available along with the code at the project repository at https://github.com/wise-ch/wastewater-influenza-dashboard.

https://github.com/wise-ch/wastewater-influenza-dashboard

## Acknowledgments

We are grateful to the many individuals who helped with this project. Taru Singhal provided code upon which the dashboard shiny app is based. Adrian Lison helped set up and maintain the dashboard. Charlie Gan, Franziska Böni, Laura Brülisauer, Camila Morales Undurraga, Johannes Rusch, and Lea Caduff processed wastewater samples. We thank the employees of the wastewater treatment plants ProRheno AG (Basel), ARA Werdhölzli (Zurich), and STEP d’Aïre (Geneva), for providing samples. Finally, the Swiss Federal Office of Public Health provided confirmed case data that was reported under the obligatory reporting system in Switzerland. We gratefully acknowledge funding support from the Swiss Federal Office of Public Health and a Swiss National Science Foundation Sinergia grant number CRSII5_205933.

