## Supplement for "Influenza transmission dynamics quantified from wastewater"


| | |
|---|---|
| Supplemental methods | 2 |
| Supplemental figures | 3 |
| Supplemental tables | 11 |

#### Supplemental methods

For the delay between infection and respiratory shedding and the delay between infection and symptoms, we used the distributions given in Carrat et al. (2008). We assumed that this second distribution (infection to symptoms) is a good approximation for the delay between infection and an individual getting a diagnostic test. To generate gamma distributions from the data presented in Carrat et al. (2008), we digitized Figure 2 from that paper using the online tool https://automeris.io/WebPlotDigitizer/. We then matched a gamma distribution to the digitized data by calculating the mean and standard deviation of the empirical distributions of the two types of data. We used these means and standard deviations for our gamma delay distributions. Figure S3A and S3C show the correspondence between the Carrat et al. (2008) data and the fitted distributions.

For the delay between infection and fecal shedding we collated data from two previously published studies (Hirose et al. 2016; Chan et al. 2011). These included at least one patient with IBV infection, but were primarily from patients with IAV infection. We shifted the data by two days to account for the fact that measurement days were relative to symptom onset, based on an average delay from infection to symptom onset of 2 days given in Carrat et al. (2008). We calculated the mean viral concentration by day and matched a gamma distribution to these values using the moments of the empirical data distribution as above. Figure S3B shows the correspondence between the data and the fitted distribution.

#### Supplemental figures

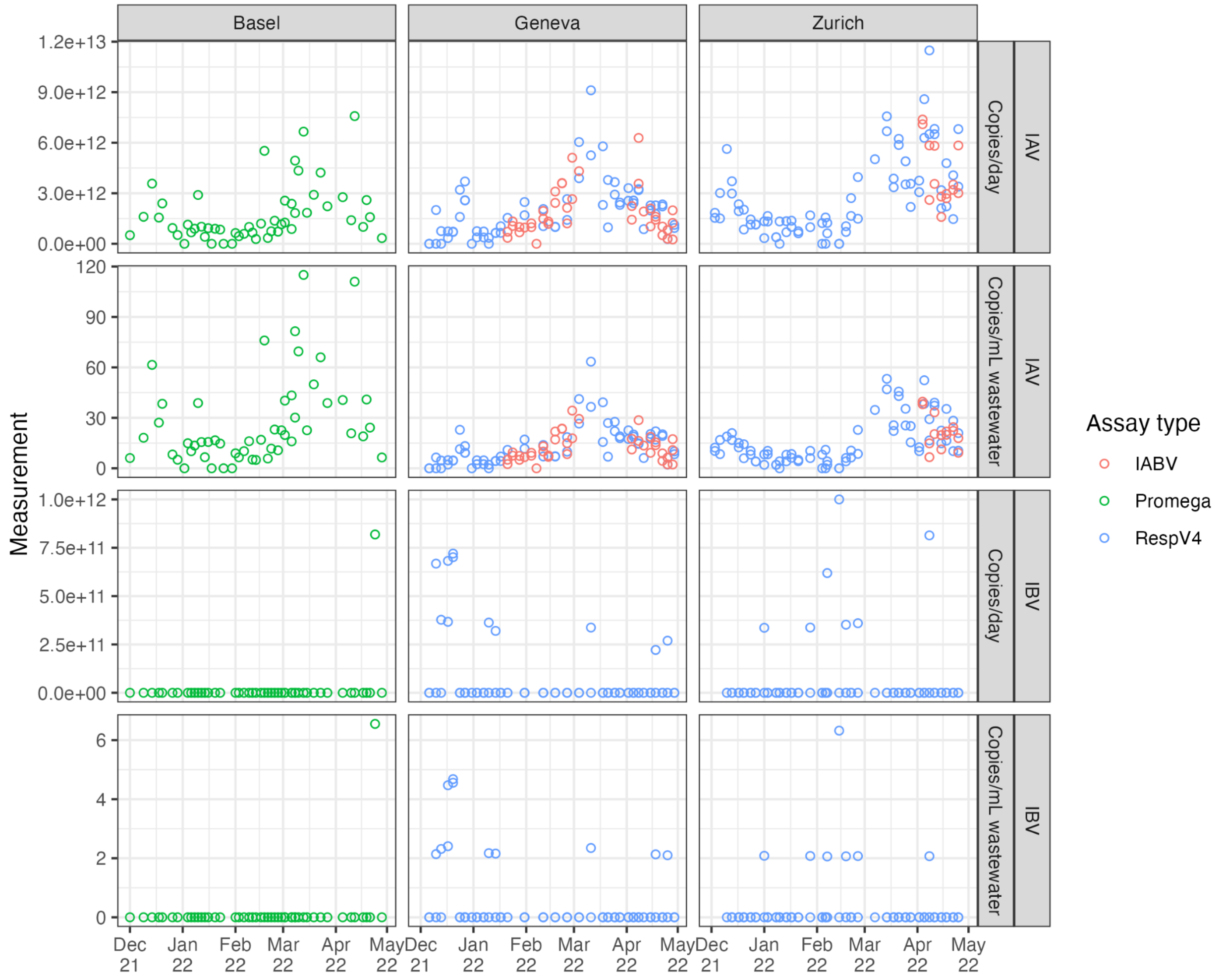


**Figure S1. Raw wastewater influenza measurements.** Each open point represents a unique measurement. Measurements from the same catchment on the same day are replicate measurements based on multiple aliquots. We use data generated using three different assays, which are shown as different colors.

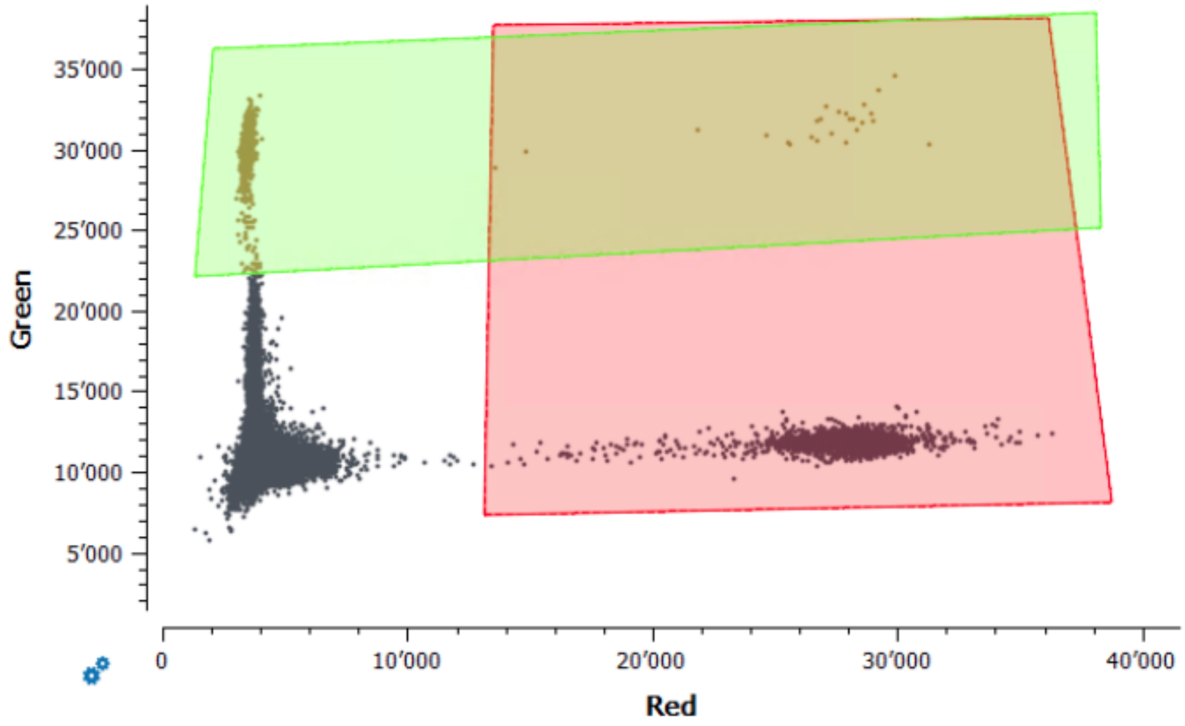


**Figure S2. Gating of positive control samples for the IABV assay.** The green fluorophore (y-axis) is for IBV and the red fluorophore (x-axis) is for IAV. The rectangles show the fluorescence thresholds used to determine which droplets in samples were positive for each virus. This assay was only used to quantify IAV because there is not a clear separation between positive and negative droplets for the IBV control.

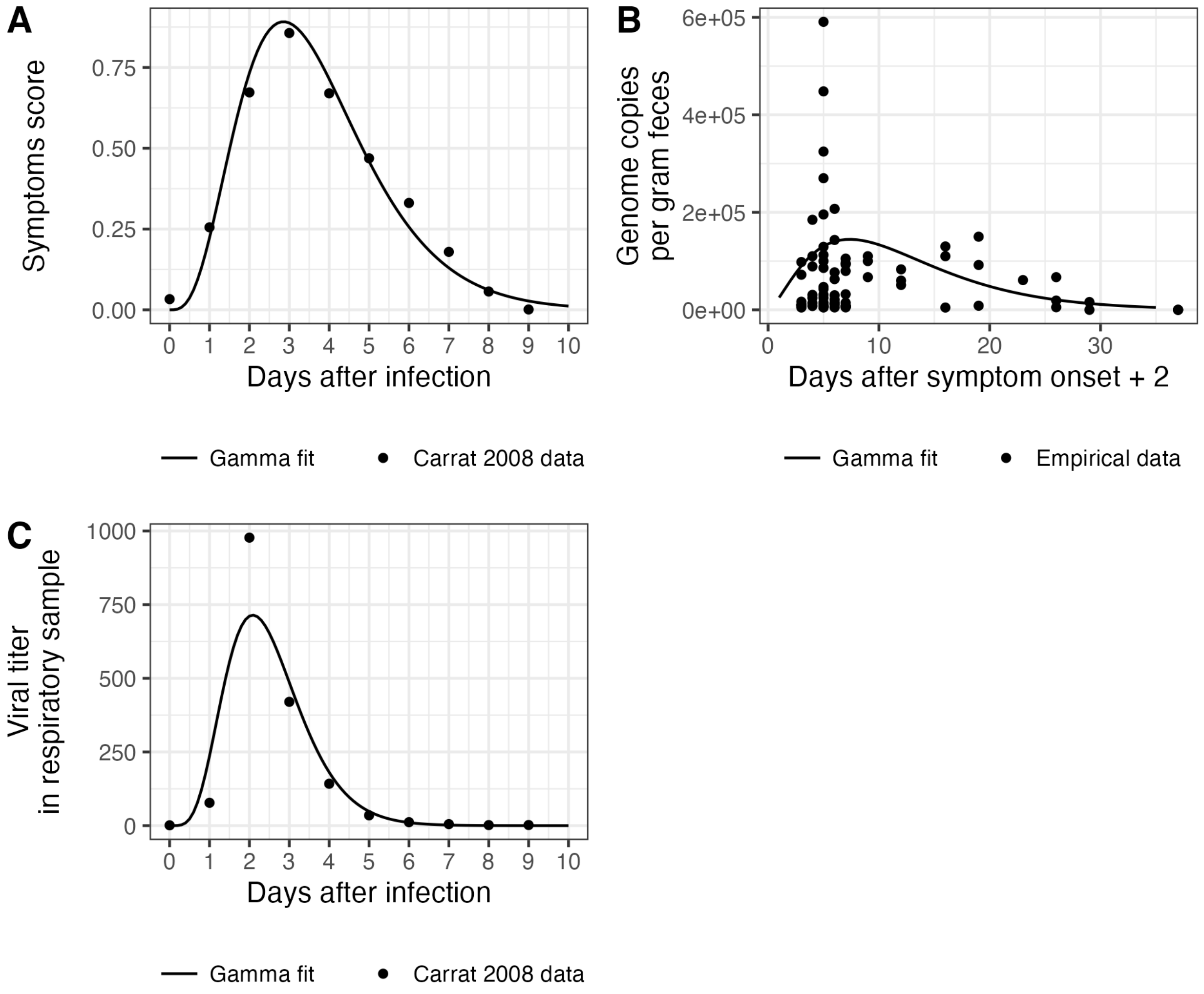


**Figure S3. Delay distributions used to reconstruct infection incidence. (A)** shows the delay distribution used for infection to a diagnostic test. **(B)** shows the fecal shedding load distribution. Measurements were taken with reference to the date of symptom onset, so here they are shifted two days to account for an assumed 2-day fixed delay between infection and symptom onset. One outlier measurement of 4639335 is not shown in this plot but was used for fitting the gamma distribution. **(C)** shows the respiratory shedding load distribution.

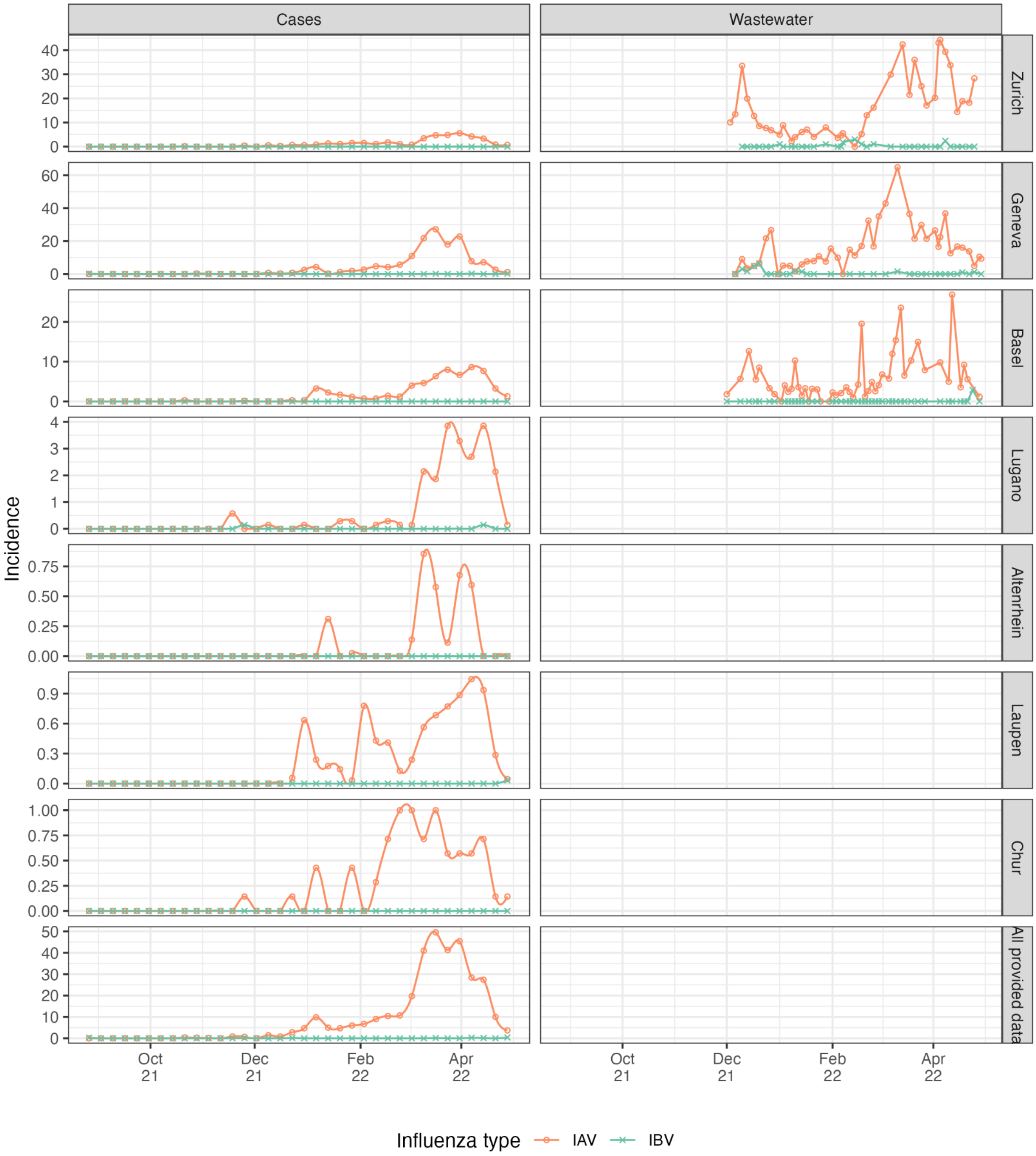


**Figure S4. Input data for Re estimation.** Cases are laboratory-confirmed influenza infections (by catchment area, reported weekly). Wastewater input data is virus load (genome copies per day) scaled by the minimum non-zero measurement for the respective wastewater treatment plant. The points show the real data and the lines show the cubic spline interpolation used to generate daily values for Re estimation.

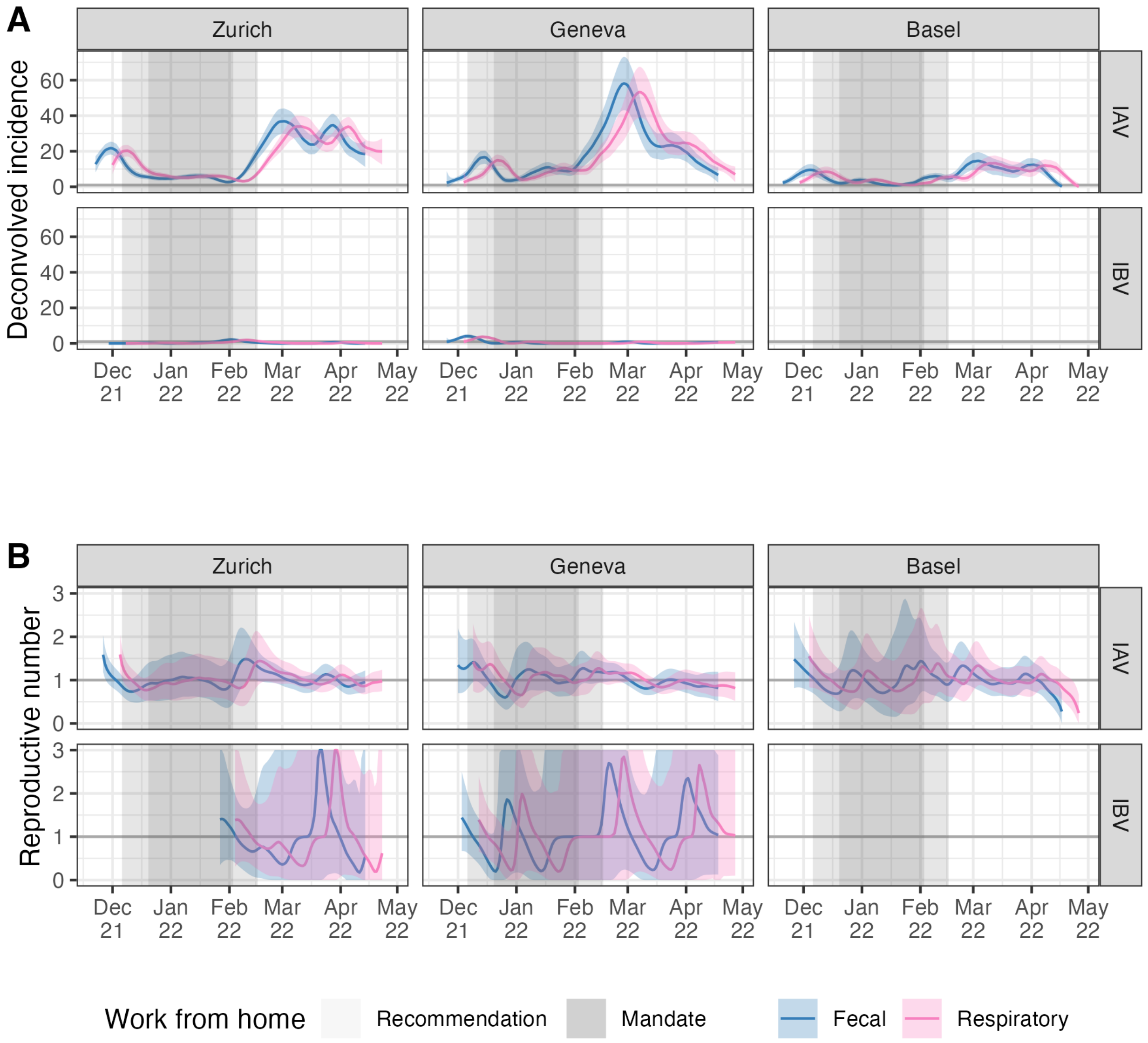


**Figure S5. Sensitivity analysis for the delay distribution.** Here we used two different shedding load distributions to deconvolve wastewater influenza measurements to infection incidence, one based on measurements of influenza in stool ("Fecal") and one based on measurements of influenza shedding from the respiratory tract ("Respiratory"). The two distributions are shown in Figure S3. **(A)** shows the infection incidence estimates using each distribution and **(B)** shows the reproductive number estimates. The thicker gray horizontal line in (B) shows the epidemic threshold at a reproductive number of 1. The shaded gray rectangles highlight periods when work from home measures were in place.

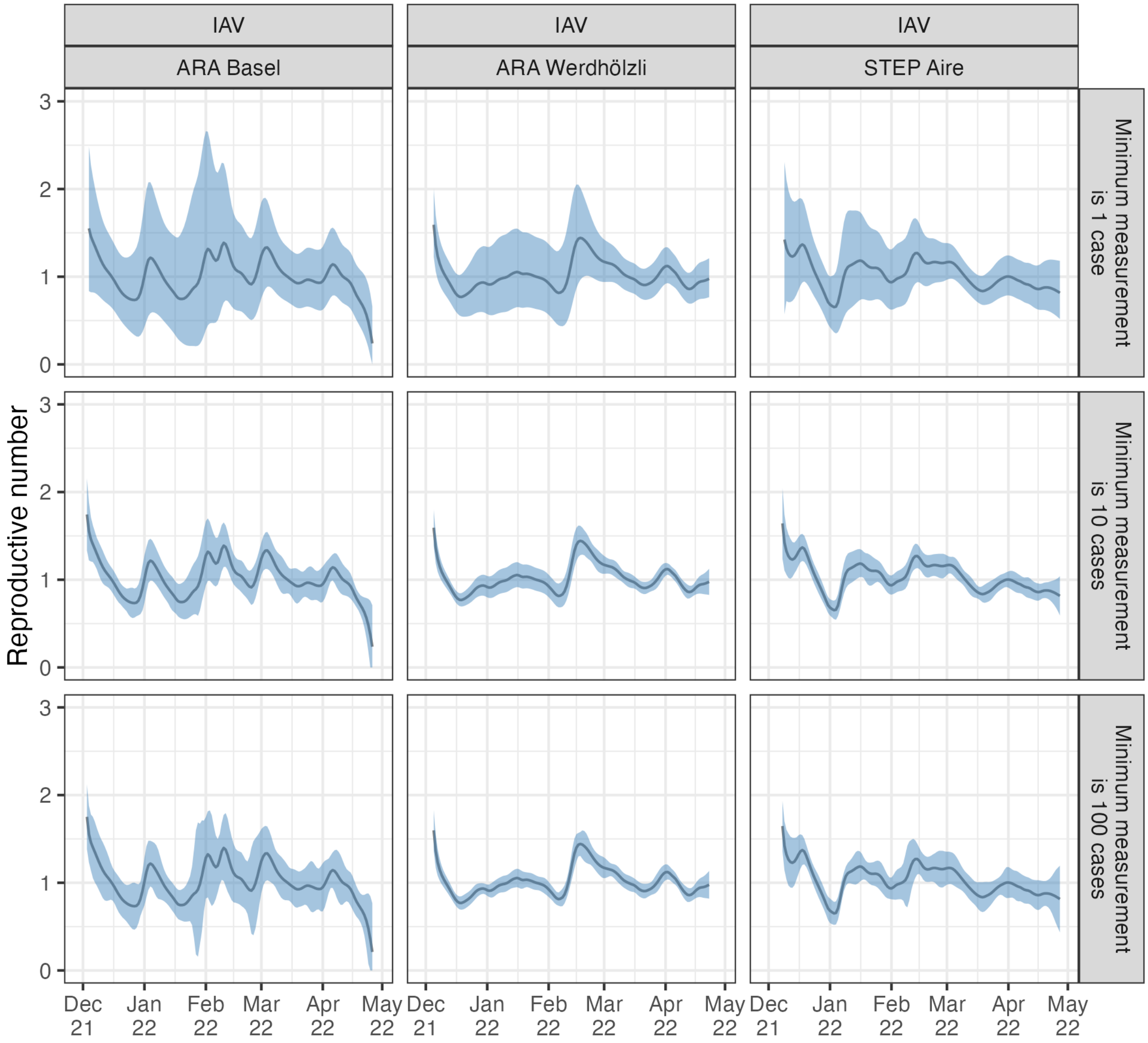


**Figure S6. Sensitivity analysis for wastewater measurement scaling.** Results are based on the same wastewater loads for IAV from the Zurich catchment, but scaled so that the minimum detected load corresponds to 1, 10, 100, or 1000 assumed cases in the catchment area.

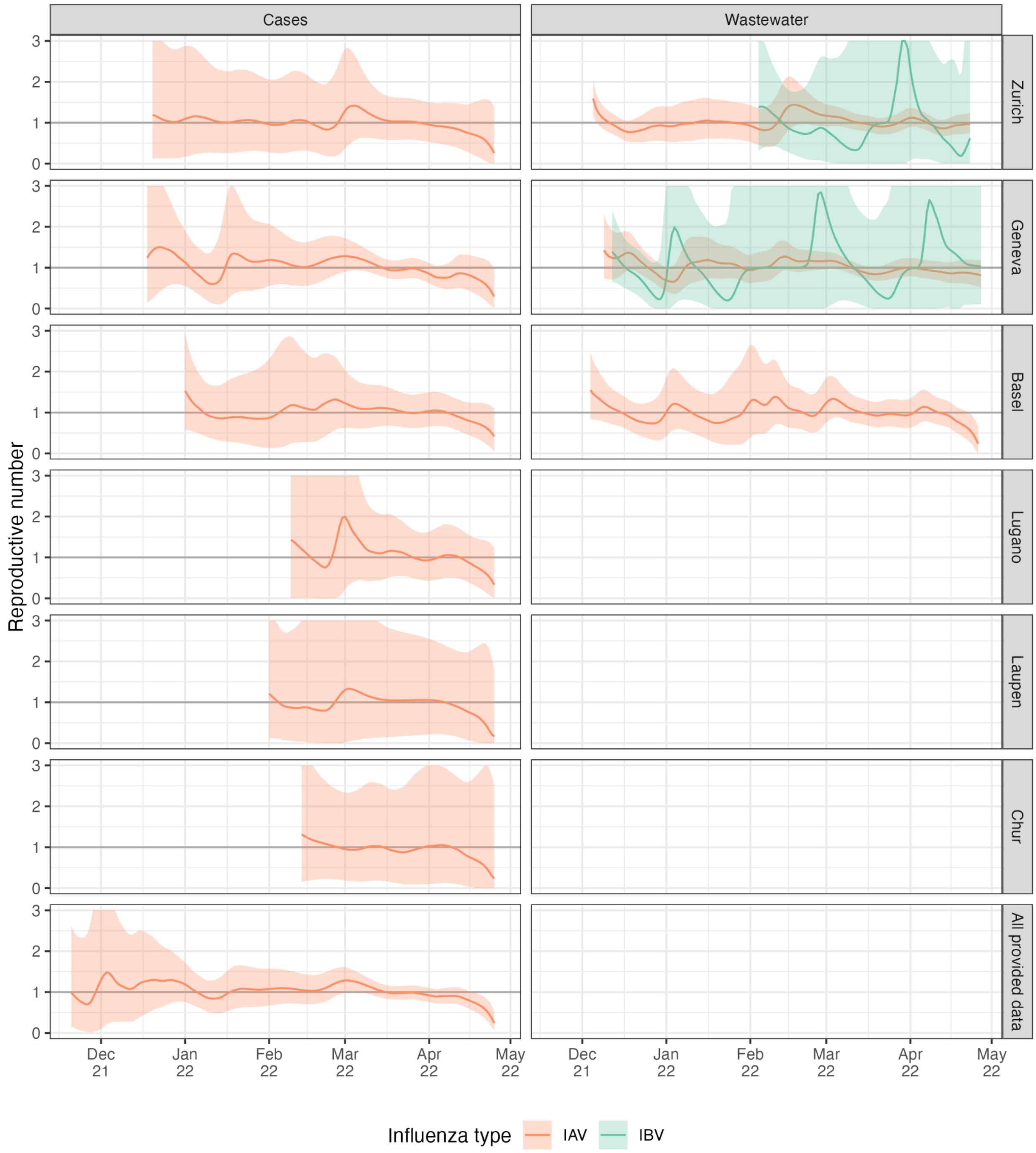


**Figure S7. Reproductive number estimates for 6 different catchment areas and all postal codes for which we have case data.** Results are not output when the cumulative number of deconvolved cases is less than 12, which is the case for influenza B virus for most case-based data and for influenza A virus in the case data from the Altenrhein catchment area. The thicker gray horizontal line shows the epidemic threshold at a reproductive number of 1.

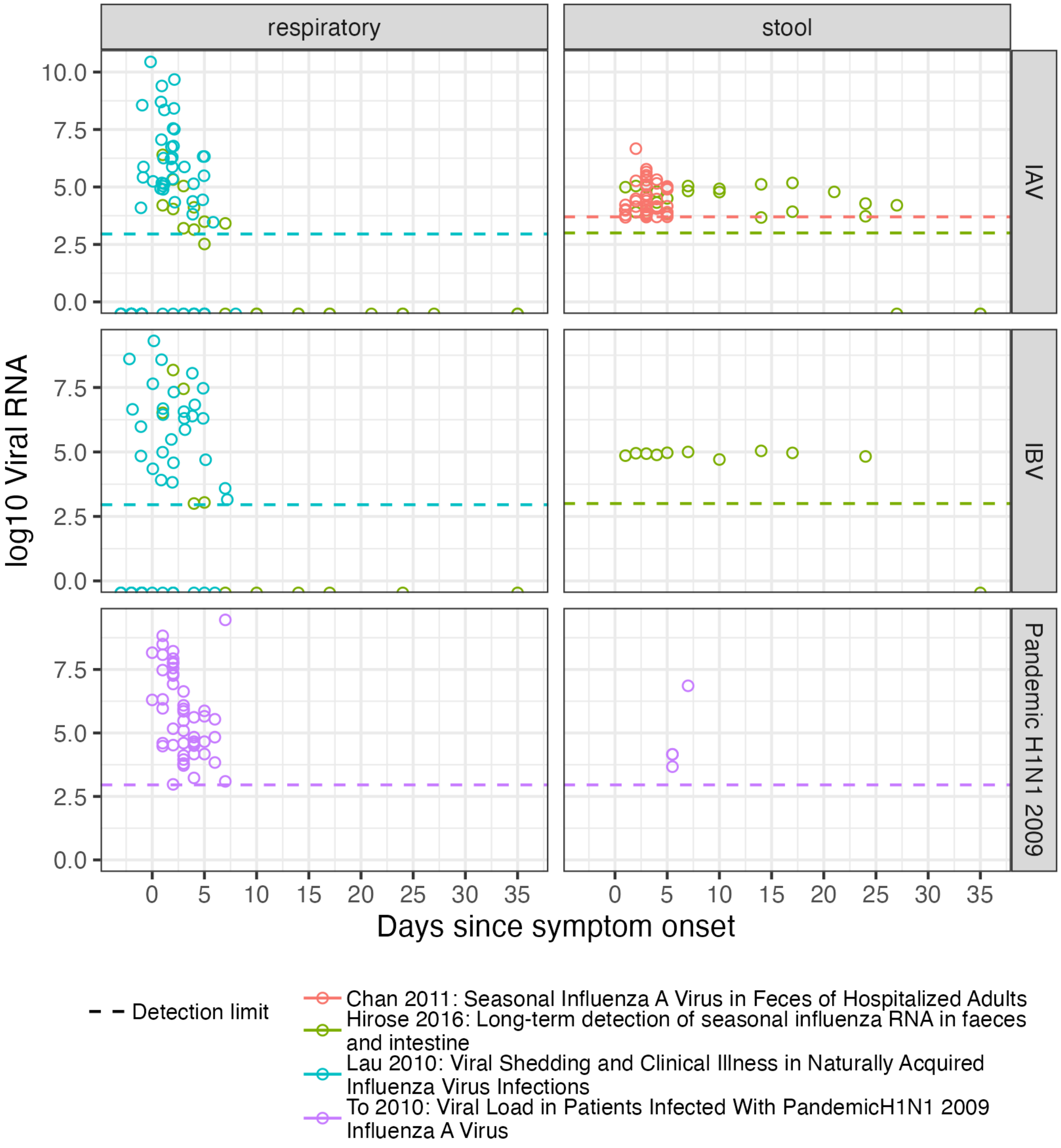


**Figure S8. Influenza shedding measurements from a convenience sample of previous studies.** "Respiratory" samples include nose-throat swabs and sputum. "Stool" samples are feces. Referenced studies are (Chan et al. 2011; Hirose et al. 2016; Lau et al. 2010; To et al. 2010).

### Supplemental tables

**Table S1: Primer and probe sequences.** Primers and probes shown were used in the duplex (IABV) and tetraplex (RESPV4) assays for Zurich and Geneva wastewater treatment facilities. Sequences are in the 5' -> 3' direction

| Target | Name | Sequence (5' → 3') | Sense (+/-) | Amplicon Size (bp) | Final Assay Conc. (nM) | Reference |
|---|---|---|---|---|---|---|
| Influenza A (IABV, RESPV4) Matrix protein gene (M) | IAV-M_Neo_Fwd | TGG AAT GGC TAA AGA CAA GAC CAA T | + | 89 | 500 | Modified from Ward et al. 2004 |
| | IAV-M_Neo_Rev | AAA GCG TCT ACG CTG CAG TCC | - | | 500 | |
| | IAV-M_Neo_Probe | Cy5/TTT GTK TTC ACG CTC ACC GTG CCC/BHQ2 | + | | 200 | |
| Influenza B (IABV, RESPV4) Matrix protein gene (M) | IBV-M_Neo_Fwd | GAG ACA CAA TTG CCT ACY TGC TT | + | 96 | 500 | Modified from Ward et al. 2004 |
| | IBV-M_Neo_Rev | ATT CTT TCC CAC CRA ACC AAC A | - | | 500 | |
| | IBV-M_Neo_Probe | HEX/AGA AGA TGG AGA AGG CAA AGC AGA ACT AGC/BHQ2 | + | | 200 | |
| SARS-CoV-2 (RESPV4) Nucleoprotein gene locus 2 (N2) | 2019-nCoV-N2-F | TTA CAA ACA TTG GCC GCA AA | + | 67 | 500 | SARS-CoV-2 RUO qPCR Primer & Probe Kit (IDT) |
| | 2019-nCoV-N2-R | GCG CGA CAT TCC GAA GAA | - | | 500 | |
| | 2019-nCoV-N2-P | FAM/ACA ATT TGC CCC CAG CGC TTC AG/BHQ1 | +? | | 125 | |
| Respiratory Syncytial Virus (RESPV4) Matrix protein gene (M) | RSV_Metavir_Fwd | GGC AAA TAT GGA AAC ATA CGT GAA | + | 117 | 500 | Modified from Fry et al. (2010) |
| | RSV_Metavir_Rev | GGC ACC CAT ATT GTW AGT GAT G | - | | 500 | |
| | RSV_Metavir_Probe | ATTO620/GCT GTG TAT GTG GAG CCY TCG TGA AG/BHQ2 | - | | 400 | |
